# The iHealth-T2D study: Statistical analysis plan for a cluster randomised controlled trial with intensive family-based lifestyle modification programme to reduce type 2 diabetes risk amongst South Asians

**DOI:** 10.1101/2020.11.12.20229849

**Authors:** Mirthe Muilwijk, Marie Loh, Sara Mahmood, Saranya Palaniswamy, Samreen Siddiqui, Wnurinham Silva, Gary S. Frost, Heather M. Gage, Marjo-Riitta Jarvelin, Ravindra P. Rannan-Eliya, Sajjad Ahmed, Sujeet Jha, Anuradhani Kasturiratne, Prasad Katulanda, Khadija I. Khawaja, Jaspal S. Kooner, Ananda R. Wickremasinghe, Irene G.M. van Valkengoed, John C. Chambers

**Affiliations:** Amsterdam UMC, University of Amsterdam, Department of Public Health, Amsterdam Public Health research institute, Meibergdreef 9, Amsterdam, The Netherlands; Lee Kong Chian School of Medicine, Nanyang Technological University, Singapore 308232, Singapore; Imperial College London, Department of Epidemiology and Biostatistics, School of Public Health, Imperial College London, St Mary’s Campus, Norfolk Place, London W2 1PG, UK; Services Institute of Medical Sciences, Department of Endocrinology & Metabolism, Services Institute of Medical Sciences, Services Hospital, Ghaus ul Azam, Jail Road 54700, Lahore, Pakistan; University of Oulu, Center for Life Course Health Research, Faculty of Medicine, University of Oulu, Oulu, Finland; Max Healthcare, Institute of Endocrinology, Diabetes and Metabolism, Max Super Speciality Hospital, 2, Press Enclave Road, Skaet, New Delhi-110017, India; Imperial College London, Faculty of Medicine, Imperial College Hammersmith Campus, DuCane Road, London W12 ONN, UK; University of Surrey, Surrey Health Economics Centre, Department of Clinical and Experimental Medicine, University of Surrey, Leggett Building, Daphne Jackson Road, Guildford GU2 7WG, Surrey, UK; University of Oulu, Unit of Primary Care, Oulu University Hospital, Oulu, Finland; Brunel University, Department of Life Sciences, College of Health and Life Sciences, Brunel University London, Kingston Lane, Uxbridge, Middlesex UB8 3PH, UK; Institute for Health Policy, Institute for Health Policy, 72 Park Street, Colombo, 00200, Sri Lanka; Punjab Institute of Cardiology, Jail Road, Shadman, Lahore, Punjab, Pakistan; University of Kelaniya, Faculty of Medicine, University of Kelaniya, PO Box 06, Thalagolla Road, Ragama 11010, Sri Lanka; University of Colombo, Department of Clinical Medicine, Faculty of Medicine, University of Colombo, 25 Kynsey Rd, Colombo 00800, Sri Lanka; London Northwest University Healthcare NHS Trust, London Northwest University Healthcare NHS Trust, Uxbridge Road, Southall, Middlesex UB1 3HW, UK; Imperial College London, National Heart and Lung Institute, Imperial College London, Hammersmith Hospital Campus, DuCane Road, London W12 ONN, UK

**Author notes:** MM and ML contributed equally and are joint first authors. IGMV and JCC contributed equally and are joint senior authors. Corresponding author: Mirthe Muilwijk.

**Keywords:** Type 2 diabetes, South Asian, Lifestyle intervention

## Abstract

**Background:** South Asians are at high risk of type 2 diabetes (T2D). Lifestyle modification is effective at preventing T2D amongst South Asians, but the approaches to screening and intervention are limited by high-costs, poor scalability and thus low impact on T2D burden. An intensive family-based lifestyle modification programme for prevention of T2D was developed. The aim of the iHealth-T2D trial is to compare the effectiveness of this programme with usual care.

**Methods:** The iHealth-T2D trial is designed as a cluster randomised controlled trial (RCT) conducted at 120 locations across India, Pakistan, Sri Lanka and the UK. A total of 3,682 South Asian men and women with age between 40-70 years without T2D but at elevated risk for T2D [defined by central obesity (waist circumference ≥95cm in Sri Lanka, or ≥100cm in India, Pakistan and UK) and/or prediabetes (HbA1c ≥6.0%)] were included in the trial. Here we describe in detail the statistical analysis plan (SAP), which was finalised before outcomes were available to the investigators. The primary outcome will be evaluated after three years of follow-up after enrolment to the study, and is defined as T2D incidence in the intervention arm compared to usual care. Secondary outcomes are evaluated both after one and three years of follow-up and include biochemical measurements, anthropometric measurements, behavioural components and treatment compliance.

**Discussion:** The iHealth-T2D trial will provide evidence whether an intensive family-based lifestyle modification programme in South Asians who are at high risk for T2D is effective in the prevention of T2D. The data from the trial will be analysed according to this pre-specified SAP.

**Ethics and dissemination:** The trial was approved by the international review board of each participating study site. Study findings will be disseminated through peer-reviewed publications and in conference presentations.

**Trial registration:** EudraCT 2016-001350-18. Registered 14 April 2016 https://www.hra.nhs.uk/planning-and-improving-research/application-summaries/research-summaries/ihealth-t2d/; ClinicalTrials.gov NCT02949739. Registered 31 October 2016, https://clinicaltrials.gov/ct2/show/NCT02949739, First posted 31/10/2016.

## Introduction

Type 2 diabetes (T2D) is the fifth leading cause of death worldwide(1) and a major contributor to the development of various comorbidities including coronary heart disease, stroke, peripheral vascular disease and end-stage renal failure(2). South Asians, who represent one-quarter of the world’s population, are at high risk of T2D and its complications, both in the country of origin and after migration(3, 4). Key modifiable risk factors that could be targeted to delay or prevent the onset of T2D include behavioural factors such as diet and physical activity(5). In past decades evidence from “the Finnish Diabetes Prevention Study” and “the Diabetes Prevention Program” showed that targeting these behavioural factors may be effective to delay or prevent the onset of T2D(6, 7). A recent meta-analysis on 1,816 participants from six randomized controlled trials (RCTs) (four from Europe and two from India) has reported lifestyle modifications may also be effective among South Asian populations(8).

Although the studies conducted to date provide some support for the utility of lifestyle interventions for prevention of T2D amongst South Asians, there are significant limitations. First, completed studies conducted in India and Sri Lanka are limited to local settings and small sample sizes, and there are no studies reported from Pakistan. Second, the evidence-based approach established by studies to prevent T2D among South Asians lack scalability and sustainability for T2D prevention, especially in low-middle income settings, since previous lifestyle interventions to prevent T2D were designed in a way that makes them labour-intensive and costly. To address these important limitations, we designed the iHealth-T2D trial in a way that makes the intervention scalable and sustainable in both low-middle income and high-income settings. The iHealth-T2D intervention aims to identify participants as being at risk for T2D based on parameters which include low-resource strategies such as waist circumference, and to improve cost-effectiveness and scalability of lifestyle modification through use of community health workers and a family-based lifestyle modification. Furthermore, we aimed to evaluate the effectiveness of the intervention in different cultural groups living in India, Pakistan, Sri Lanka and the UK, to improve generalisability. The objective of the iHealth-T2D trial is to investigate whether our family-based lifestyle modification delivered by community health workers is effective to prevent T2D amongst South Asians at high risk for T2D (based on central obesity or pre-diabetes), compared to usual care.

Here we report the details of the statistical analysis plan (SAP), prepared according to the published guidelines on the content of SAPs(9). This SAP includes details on the analyses of the primary objective, but does not include details on secondary questions nor the evaluation of cost-effectiveness. The cluster RCT is registered with EudraCT: 2016-001350-18 and ClinicalTrials.gov: NCT02949739. This SAP should be read in conjunction with the study protocol, which contains more details on the study rationale and design. The study protocol will be published and is until then available upon reasonable request.

### Summary study design

The iHealth-T2D trial is designed as a cluster RCT amongst 3,682 South Asians at high risk for T2D at 120 locations across India, Pakistan, Sri Lanka and the UK. The study design is summarised in *Figure 1*., in brief: a total of 120 sites from a range of socio-economic settings were identified, comprising 30 sites in each of the four participating countries (India, Pakistan, Sri Lanka and the UK). The field-work sites were cluster randomised to either family-based lifestyle modification or usual care (1:1 allocation). At each field-work location we aimed to recruit 15 male and 15 female South Asians between the ages of 40 and 70 years old, at high risk, but free from T2D, were included. High risk for T2D was defined by central obesity (waist circumference ≥95cm in Sri Lanka, or ≥100cm in India, Pakistan and UK) and/or prediabetes (HbA1c≥6.0%). Exclusion criteria included participants with known type 1 or type 2 diabetes, fasting glucose levels ≥7.0 mmol/L, HbA1c levels ≥6.5%, BMI <22 kg/m^2^, pregnant or planning pregnancy, unstable residence or planning to relocate and serious illness.

**Figure 1:**
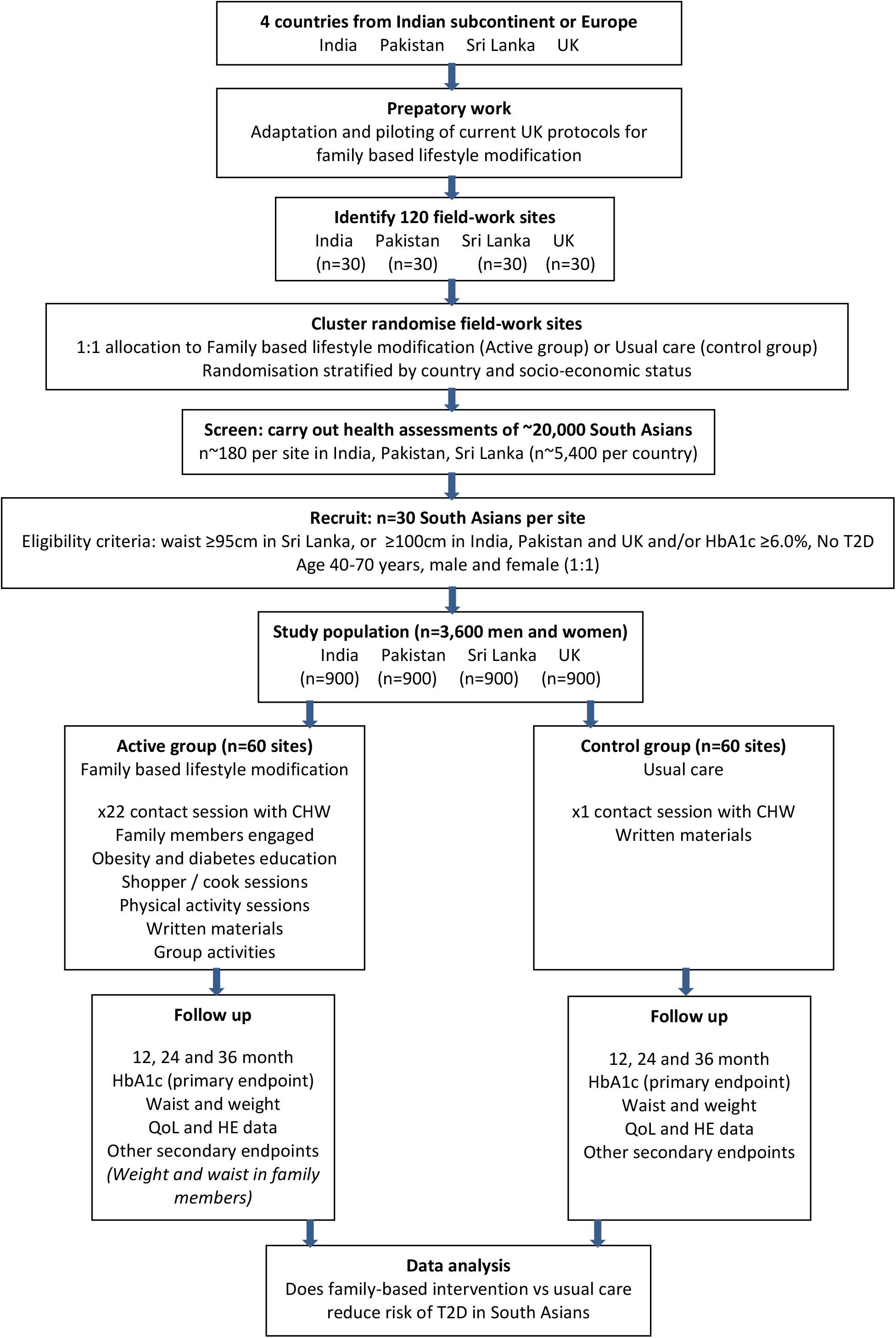
Study Flow

The primary aim of the lifestyle modification sessions was the prevention of T2D, details on the various sessions can be found in the iHealth-T2D study protocol. In brief, participants in the lifestyle modification arm received family-based lifestyle modification delivered by a community health worker, consisting of 22 contact sessions over a period of 12 months. Participants in the usual care arm received a single diabetes prevention education session lasting 30-60 minutes delivered by a community health worker. Written material was distributed additionally. Both participants and community health workers could not be blinded to the treatment arm as the type of care given is clearly visible. They were, however, kept masked to outcome measurements and trial results. Participants were followed up annually, during a period of three years. Data were obtained by research nurses who were blinded to trial arms to reduce the risk of assessment bias.

Ethical approval was obtained from the Institutional Review Board in each participating country and at each research location before the start of the study. Information sheets and consent forms were made available in the major South Asian languages. Multilingual translators were available as required. Each participant provided informed consent. People unwilling or unable to provide consent were excluded from the study. The research complied with relevant national and international regulations and was carried out in line with the Declaration of Helsinki and the International Conference on Harmonization Guidelines.

### Study outcomes

The effectiveness of the iHealth-T2D intervention will be evaluated at two time points, both after one and after three years of follow-up. After one year of follow-up secondary outcomes are evaluated, while after three years of follow-up both the primary outcome and secondary outcomes will be evaluated.

The primary outcome is defined as: T2D incidence in the intervention arm compared to the usual care arm after three years of follow-up in both groups of South Asians with central obesity and prediabetes. T2D incidence is defined as a physician diagnosis and being on treatment for T2D, or HbA1c levels ≥6.5% (10). The primary outcome will not be evaluated after one year of follow-up as power calculations were based on a three year follow-up duration.

Secondary outcomes include glucose, insulin, HOMA, total cholesterol, HDL cholesterol, triglycerides, blood pressure, waist circumference, weight, smoking status, alcohol use, physical activity, dietary intake, treatment compliance and dose delivered / received, are, therefore, evaluated both after one and three years of follow-up. The evaluation of secondary outcomes after one as well as three years of follow-up will allow for comparisons of both short- and long-term effects of the lifestyle intervention. In addition, subgroup analyses amongst participants included in the study based on HbA1c and/or waist circumference will be performed.

Study outcomes that will be reported but are not included in the current SAP are secondary questions which include changes in adiposity and glucose homeostasis amongst the extended family, psychosocial measures, cost effectiveness and implications of scaling up locally and nationally.

Power calculations performed before the start of the data collection in the first version of the trial protocol showed that the iHealth-T2D trial is well powered to assess the primary outcome *(Additional file 1)*. These a priori power calculation considered adjustment for multiple testing (a *p*<0.017), because the effects in the main analysis were calculated by subgroups based on a high risk for T2D either based on HbA1c and/or waist circumference. This initial protocol and the subsequent power calculations were, however, amended.

In the revised protocol, as reported here, the primary study outcome assessment is based on the overall effects in the total group, with the analyses by subgroup reported as supplementary information. This means that the main analyses will not be adjusted for multiple testing, which is in line with current recommendations(11). Additionally, the revised power calculations now include a variation inflation factor, also known as the design effect, to take the cluster randomisation into account. For the power calculations, we assumed an intraclass correlation coefficient (ICC) of 0.01. This was based on prior observations that intent-to-change behaviours related to diet and physical activity which are generally not very strongly clustered. The findings in previous work report that median ICCs in primary care lay around 0.01(12, 13). This may overestimate the impact, as inclusion of baseline co-variates, as planned in the current protocol, have been shown to reduce the design effect, e.g. to a median of 0.005 for ICCs in primary care(13, 14). Nevertheless, such a correction reduces the power of the study as compared to analyses that do not account for design effects.

With this new focus and design effect the power of the study can be calculated as follows. Assuming that event rates for T2D for usual care are 6.8% per year(15), the study has 81% power to identify at *p*<0.05 a reduction in T2D incidence of 35%(8), after a follow-up period of three years with an estimated drop-out rate of 10%. The study was, thus, sufficiently powered for the primary outcome. Since secondary outcomes are of continuous nature, it is expected that a statistically significant change can already be detected after one year of follow-up *(Table 1)*.

**Table 1:** Detectable difference in secondary study outcomes after one year of follow-up.

|  |  | Detectable difference |  |  |
| --- | --- | --- | --- | --- |
| Measure | Expected SD | 0% Dropout | 10% Dropout | 20% Dropout |
| Waist (cm) | 11.5 | 1.23 | 1.28 | 1.34 |
| Weight (kg) | 7.5 | 0.80 | 0.84 | 0.88 |
| Glucose (mmol/L) | 0.6 | 0.06 | 0.07 | 0.07 |
| HbA1c (%) | 0.5 | 0.05 | 0.06 | 0.06 |
| HOMA | 3.0 | 0.32 | 0.33 | 0.35 |
| Total cholesterol<br>(mmol/L) | 0.9 | 0.10 | 0.10 | 0.11 |
| Triglycerides (mg/dL) | 1.9 | 0.20 | 0.21 | 0.22 |
| Blood pressure systolic<br>(mmHg) | 16 | 1.71 | 1.78 | 1.87 |
| Blood pressure diastolic<br>(mmHg) | 8.6 | 0.92 | 0.96 | 1.00 |
| Dietary intake (Kcal) | 390 | 41.7 | 43.4 | 45.5 |

*Expected detectable difference for a range of secondary study outcomes after 1 year of follow-up, assuming i)* *standard deviation (SD) of 10 cm with a two-sided 5% significance level and ii) ICC of 0.01. iii) a two sided p-* *value of 0.05 iv) a power of 0.80.*

### Statistical analysis plan

#### General principles

The analysis of the primary outcome will be performed after three years of follow-up. Secondary outcomes will be evaluated both after one and three years of follow-up. Analyses will be performed by investigators of the iHealth-T2D study group (MM for the 1-year analyses), who were blinded for the intervention, on a clean anonymised data set (JCC and AK). The latest version of the R statistical software package will be used. Tests will be two-sided and *p*-values <0.05 will be considered statistically significant. We will not adjust for multiple testing as we pre-defined primary and secondary outcomes (11). Data will be reported in line with the Consolidated Standards of Reporting Trials (CONSORT) 2010 Statement (16).

The analyses will be performed according to the intention-to-treat (ITT) principle with data from all participants enrolled in the study (17). Data of participants who attended at least one post-baseline assessment will be analysed according to their initially assigned study arm, regardless of their adherence. Participants who withdrew their consent will be excluded from the ITT analyses, the number of participants who withdrew consent and reasons for withdrawing consent will be reported. The patterns of missing data for primary and secondary outcomes and, if known, reasons for missingness, will be summarised for both treatment arms (18). The nature and pattern of missing data will be explored. If data missing at random is assumed, data will be imputed by multiple imputation methods, if data is not missing at random a ‘best case/worst case’ sensitivity analysis will be used (19). In case multiple imputation is used for one or more outcomes, variables selected as predictors for imputation contain known predictors of T2D. These include co-variates included in the main analysis model (sex, age) and the auxiliary variables country, setting, socio-economic status, pack-years of smoking, alcohol consumption, metabolic equivalents (METs) of physical activity, waist circumference and HbA1c. Missing values will be imputed separately by allocated randomisation group (20), and will comply with the multi-level character of the data.

All statistical analyses will be adjusted for confounders registered at baseline, namely age and sex.

#### Baseline characteristics

Baseline characteristics of both study arms will be presented by sex and country and presented in a table. The baseline characteristics will not be tested for statistical differences between study arms (21). The baseline characteristics will be reported by arithmetic means and standard deviation (normally distributed numerical data), medians and interquartile ranges (non-normally distributed numerical data) or percentages and numbers (categorical data). Normality of data distributions will be inspected visually by plotting histograms and we will assess the deviation from normality by the Shapiro-Wilks test. In case of a *p*-value >0.05 the data will be transformed for normality.

Descriptive characteristics to report at baseline include: age (years), setting (%), socio-economic status (%), smoking (pack-years), alcohol consumption (units/week), physical activity (MET/week), BMI (kg/m^2^), waist circumference (cm), HbA1c (%), and glucose (mmol/L). The descriptive characteristics will be reported by treatment arm. The population size and number of missing observations will also be reported.

#### Analyses of primary outcome

Cumulative incidence of T2D will be summarized and compared between treatment arms using random effects logistic regression to estimate odds ratios (OR) and 95% CI. In addition we will evaluate intraclass correlation coefficients to assess cluster variance. The model will include the randomisation stratum site as a random effect and country as a fixed effect. The effectiveness of the lifestyle intervention will be reported by the screening numbers needed to identify one case of ‘high risk’ for developing diabetes and the number needed to treat or delay one case of T2D. The Wilson score method will be used to calculate CIs (22).

#### Analyses of secondary outcomes

The secondary outcomes are of continuous nature and will be reported as mean and SD in each of the two treatment groups. The differences between the two treatment arms will be estimated with a multilevel linear mixed-effects regression model. The models will include the stratification variable site as a random effect and country as a fixed effect to adjust for potential cluster differences. The estimates will be presented with their associated 95% confidence intervals (CIs) and *p*-values for comparison between the treatment groups. In addition, adjustment for age and sex will be performed and the baseline values will be reported.

#### Additional analyses

Treatment compliance will be reported for the intervention arm as an explanatory variable. It will be reported according to the number of times a participant turned up for the lifestyle modification (LSM) sessions. In addition, change in dietary intake will be reported. 24-h dietary recalls and food-frequency questionnaires were performed in the treatment arm only. Dietary variables will, therefore, be reported as change from baseline in the treatment arm.

Both absolute and relative risk reduction will be compared for subgroups of participants included in the study based upon a high risk for T2D according to waist circumference measurements and those included based upon HbA1c levels. The interaction of treatment arm with sex, setting, socio-economic status, baseline waist circumference and HbA1c levels will be assessed. If there is an interaction, effect estimate and p-values will be presented by subgroups.

Sensitivity analyses will be performed to identify potentially extreme centres, because extreme deviation of one site from other sites may have a large impact on the overall results.

Since lifestyle interventions are generally considered to be safe, no (serious) adverse events are to be expected. In case of any adverse events these will be reported per incident with the number per group and a description of the event.

## Discussion

The iHealth-T2D cluster RCT will provide evidence whether an intensive family-based lifestyle modification programme delivered by community health workers compared to usual care is effective to prevent T2D amongst South Asians at high risk for T2D based on central obesity or pre-diabetes, and living in India, Pakistan, Sri Lanka and the UK. Here we have provided details of the planned statistical analyses of the iHealth-T2D cluster RCT and pre-specified primary and secondary outcomes, both after one and three years of follow-up, together with planned analyses. Statistical decisions may influence final conclusions of the intervention’s effectiveness. Reporting the SAP before commence of analyses increases transparency (9) and reduces the risk of bias by outcome reporting and data-driven analyses (23). This SAP contains details on all elements of the statistical analyses limiting the risk of bias, e.g. by reporting only outcomes for which a statically significant effect was identified (23).

Although statistical methods are chosen as objectively as possible, there is always unavoidable subjectivity involved. In addition, multiple perspectives may be relevant to each statistical discussion which may lead to different choices. In addition, chosen thresholds may be somewhat arbitrary. An example is the common use of *p*-values which is currently under debate (24). We will report *p*-values and label <0.05 as statistically significant, but will also consider effect sizes to help identify the effectiveness of the trial. Reporting a statistical analysis plan will help to counteract selection bias considering not reporting study results above the *p*-value threshold, since it promotes publication of non-significant findings (25). This will make the cumulative evidence of multiple individual studies more reliable.

The analyses will be based on ITT, which is currently considered as the golden standard for RCTs (17). In ITT based analyses subjects that did not comply with the assigned treatment of the study arm or dropped out of the study are still included in the analyses according to their assigned study arm. A drawback of this approach is that the effect size of the treatment will thus be underestimated, and results may be more susceptible to type II errors. In addition, interpretability might become difficult since dose-response is unclear. An advantage of the ITT is that participants that are less likely to comply with the intervention and are thus more likely to drop out are still included in the study results. The ITT approach will thus give study results that take likeliness of adopting the lifestyle intervention in the general population into account. Altogether, the analyses based on ITT will be conservative, but mostly unbiased since balance in participants generated by the random treatment allocation is maintained.

Multiple imputation will be used in case data missing at random can be assumed. However, none of the statistical techniques currently available can completely compensate for the lack of true data. In addition, the estimates of treatment effect only remain unbiased in case the analysis model is correctly specified (20). Bias is minimised if imputation is carried out separately by randomisation group. This approach may, however, be less conservative. Participants with missing data are more likely to be non-compliant with the lifestyle intervention, while imputed data may reflect those without missing data in the treatment arm. Other approaches to deal with missing data include last observation carried forward and complete-case analysis, but both are sensitive to generate biased estimates, and we therefore did not consider these approaches (26).

In conclusion, providing the details of the SAP for the iHealth-T2D trial will help to minimise bias in publication of our study outcomes. The selected statistical methods were based on current consensus on the most appropriate methods according to scientific literature, but are always under debate. Sensitivity analyses will, therefore, include conservative estimates of the effect of the iHealth-T2D trial. If the iHealth-T2D intervention is proven effective, this family-based lifestyle modification is designed in a way that it may be used in a wide-range of settings, including those with a low availability of resources, to prevent T2D among South Asians.

### Trial status

Version: 1.0 Date: February 7, 2020

This document has been written based on information contained in the Clinical Study Protocol version 2, dated 01/11/2016.

### SAP revision history

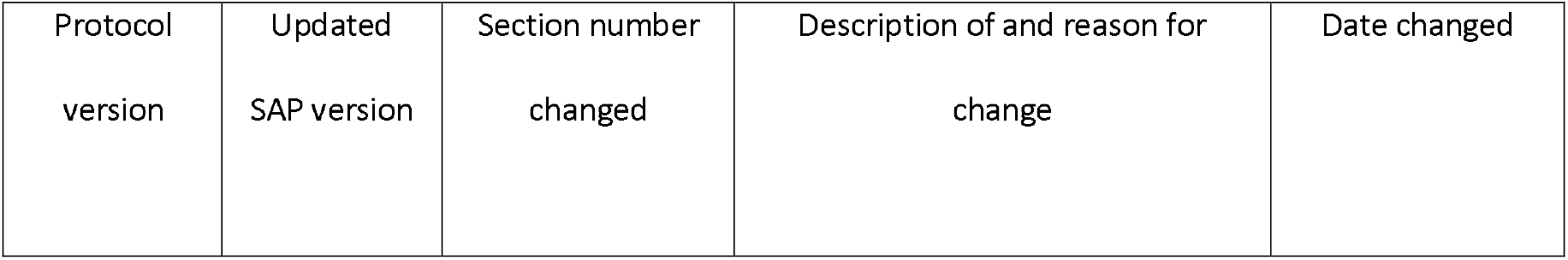

The first participant was enrolled at 15/06/2016 and the last participant at 05/03/2019. The last scheduled follow-up data is 05/03/2022.

## Data Availability

Data will be available to others on completion of the research, by application to the Steering Committee.

## List of abbreviations

CI: Confidence interval
CONSORT: Consolidated Standards of Reporting Trials
CRT: Controlled randomized trial
ICC: Intraclass Correlation Coefficient
ITT: Intention to treat
LSM: Lifestyle modification
MET: Metabolic equivalents
OR: Odds ratio
SAP: Statistical Analysis Plan
SD: Standard deviation
T2D: Type 2 diabetes

## Declarations

### Ethics approval and consent to participate

Ethical approval was obtained from the Institutional Review Board in each participating country and at each research location before the start of the study. Information sheets and consent forms were made available in the major South Asian languages. Multilingual translators were available as required. Each participant provided informed consent. People unwilling or unable to provide consent were excluded from the study.

### India

- Max Healthcare Institutional Ethic Committee (ref: CT/MSSH/SKT-2/ENDO/IEC/14-40, date 22/04/16)
- Indian Council for Medical Research (ICMR, ref: 55/7/Indo-Foreign-Diab/2014-NCD-II, date 08/02/2016)

### Pakistan

- Punjab Institute of Cardiology Ethical Committee (ref: rtpgme-research-047, date 09/04/16)
- Services Institute of Medical Sciences Institutional Review Board (ref: IRB/2016/222/SIMS, date 12/03/16)

### Sri Lanka

- University of Colombo Ethics Review Committee (ref: EC-16-063, date 23/05/16)
- University of Kelaniya Ethics Review Committee (ref: P/62/05/2016, date 11/05/16)

### UK

- West Midlands - Solihull Research Ethics Committee (ref: 16/WM/0171, date 14/04/2016)

### Consent for publication

Not applicable, no datasets are included in this SAP.

### Authors’ contributions

MM drafted and edited the manuscript. SP and IGMV reviewed and edited the manuscript. GSF, HMG, MRJ, RPRE, SA, SJ, AK, PK, KIK, JSK, RW, and JCC designed the iHealth-T2D trial. All authors reviewed the manuscript. All authors read and approved the final manuscript.

## Acknowledgements

We are most grateful to the participants of the iHealth-T2D study, and all the staff members who have taken part in gathering the data of this study.

